# Exploring the Relationship Between Education and Academic Ability in Childhood with Healthcare Utilisation in Adulthood: Findings from the Aberdeen Children of the 1950’s (ACONF)

**DOI:** 10.1101/2024.10.08.24315078

**Authors:** S Stannard, SDS Fraser, R Owen, A Berrington, S Paranjothy, NA Alwan

## Abstract

**Background:** Interactions with secondary care, including multiple outpatient appointments and hospital admissions, represents a common and often burdensome aspect of healthcare utilisation for people living with multiple long-term conditions. Lifecourse factors such as education and academic ability may play a role in shaping the risk of healthcare utilisation later in adulthood. We explored the association between education and academic ability in childhood and both outpatient appointments and hospital admissions in adulthood, accounting for the mediating role of adult factors, including long-term conditions.

**Method:** The analytical sample consisted of 7183 participants in the Aberdeen Children of the 1950’s. Three outcomes were measured using routine healthcare records (SMR00/SMR001/SMR004) over a five-year period (2004-2008) using the ‘burden’ cut-offs of: (1) ≥5 outpatient appointments, (2) ≥2 hospital admissions, or (3) ≥3 outpatient appointments plus ≥1 hospital admission. We constructed a childhood (age 6-11) education and academic ability domain and calculated predicted risk scores of the three outcomes for each cohort member. Nested logistic regression models investigate the association between domain predicted risk scores and odds of each of the three outcomes accounting for childhood confounders (maternal age, Rutter behaviour, physical grade at birth, birthweight, sex mother’s pre-marital occupation, and father’s social class) and self-reported adult mediators, including body mass index, smoking, employment status, housing tenure, having long-term conditions, and age left school.

**Result:** Adjusting for childhood confounders, lower childhood education and academic ability was associated with ≥ 5 outpatient appointments (OR1.03 95%CI 1.01-1.05), ≥ 2 hospital admissions (OR1.04 95%CI 1.03-1.6) and combined ≥3 outpatient appointments plus ≥ 1 hospital admissions (OR1.04 95%CI 1.02-1.06). Accounting for adult mediators (including long-term conditions), associations remained statistically significant, but their effect sizes were slightly reduced. When age left school was included in the final model, the association between the exposure and the combined outpatient appointments and hospital admissions (OR1.02 95%CI 1.00-1.04), ≥ 2 hospital admissions (OR1.02 95%CI 0.99-1.05) and ≥ 5 outpatient appointments (OR1.01 95%CI 0.99-1.03) were attenuated.

**Conclusions:** Education and academic ability in early life may be related to the burden of multiple hospital admissions and outpatient appointments later in life. This relationship was not fully explained by accounting for multiple long-term conditions and other potential mediating factors in adulthood. However, the age at which the participant left school seems to substantially mediate this relationship underscoring the positive impact of time spent in formal education on health during the lifecourse.

## Introduction

Multimorbidity, commonly defined as living with two or more long-term conditions (LTCs) is common, has increased in prevalence over the last 20 years, and is having a major impact on health and social care systems and people’s lives [1-4]. Multimorbidity occurs earlier in the lifecourse among people from more socioeconomically and demographically disadvantaged backgrounds [5]. A high number of outpatient appointments and hospital admissions represents one aspect of the treatment burden experienced by those living with multimorbidity, given that individuals often need regular clinical review, commonly involving multiple clinical specialties and different hospitals. A high number of hospital appointments and admissions are also associated with other aspects of burden including the fragmentation of care [6], the time and cost burden of health service utilisation [7], and in some cases these difficulties can lead to non-attendance [8,9]. A high number of outpatient attendance and hospital admissions have also been found to impact well-being, negatively affect quality of life and reduce adherence to treatment [10-12].

Previous research has demonstrated a relationship between a higher number of LTCs and more outpatient appointments and hospital admissions [10,12,13]. Research has also proved that wider demographic and socioeconomic factors in adulthood including educational level, unemployment, socioeconomic status and sex are associated with health service utilisation including outpatient appointments and hospital admissions [10,14]. Yet the relationship between factors earlier in childhood (under the age of 18) and health service utilisation has been under-explored.

Social, economic, developmental, educational and environmental experiences in childhood can have an enduring impact on outcomes across the life course. Research has demonstrated that many early life factors are related to outcomes in adulthood including, but not limited to, health, wider family health, economic circumstances, educational circumstances, crime, life satisfaction and family formations [15-23]. Given health services are facing the challenges of ageing populations, long-term prevention strategies are likely to be increasingly significant. It is therefore important to explore whether the relationship between socioeconomic factors during childhood and the burden on healthcare utilisation persists after considering the mediating role of adult factors that are on the pathway between childhood exposures and healthcare utilisation outcomes.

In previous research [24], we developed a conceptual framework to characterise the population-level domains of early-life determinants of future multimorbidity risk. Through a scoping literature and policy review, and with the support of public and patient contributors, 12 domains of early-life risk factors of future multimorbidity risk were identified [24]. These domains covered a range of social, economic, developmental, educational and environmental factors that focused on both direct and indirect factors as well as wider systemic and structural determinants of disease and health inequalities [24,25]. Subsequently we explored [25] how these 12 domains of early-life risk factors of future multimorbidity risk could be characterised across three UK cohort studies; including the Aberdeen Children of the 1950s Study (ACONF).

In this study we aimed to explore the association between childhood education and academic ability and both outpatient appointments and hospital admissions in adulthood. We additionally explored the mediating role of adult factors including body mass index (BMI), smoking status, employment status, housing tenure, presence/number of long-term conditions and the age at which individuals left school.

This work was conducted as part of the Multidisciplinary Ecosystem to study Lifecourse Determinants and Prevention of Early-onset Burdensome Multimorbidity (MELD-B) collaboration [26], which aims to identify lifecourse time periods and targets for the prevention of early-onset, burdensome multimorbidity.

## Methods

### Dataset

We used data from the Aberdeen Children of the 1950’s study (ACONF) that has followed 12,150 cohort members born in Aberdeen, Scotland, between 1950 and 1956 [27]. During primary school cohort members took reading and maths tests as part of the Aberdeen Child Development Survey (ACDS), and these test results were linked to other school and birth records including the Aberdeen Maternity and Neonatal Databank which included perinatal and social information collated throughout the course of their mother’s pregnancy and the cohort members own birth [28]. Cohort members were traced in 1999 and 7183 cohort members responded to a postal questionnaire conducted between 2001-2003. Traced responders were more likely to be women, from a higher birth social class and have a higher childhood cognition score than non-responders [28]. Traced cohort members consented to have their Scottish Medical Records (SMR) including outpatient attendance (SMR00) general/acute inpatient and day case (SMR01), and mental health general/acute inpatient and day case records (SMR04) linked to the self-reported data [27,28].

### Outcomes

Hospital admissions and outpatient appointments were measured using linked routine secondary care electronic health records including outpatient attendance (SMR00), general/acute inpatient and day case (SMR01), and mental health general/acute inpatient and day case records (SMR04) all reported over a 5-year period between 2004-2008. This period was selected to ensure the outcomes were recorded immediately following the self-reporting of adult mediators (2001-2003), to ensure the temporal ordering of variables could be established. Hospital admissions included all elective and emergency admissions apart from those related to pregnancy and birth.

The evidence on how many admissions or appointments may represent a burden to an individual is scarce, and is likely to be subjective to an individual’s own circumstances and the nature of the appointment or admission. As such, the research team discussed what may represent burden at a population level, and in consultation with our patient and public advisory board, it was decided that the following outcomes would represent healthcare utilisation burden for many people. The first outcome was the reporting of five or more separate outpatient appointments derived from the linked outpatient attendance dataset (SMR00). Secondly, we considered the reporting of two or more separate hospital admissions derived from the general/acute inpatient and day case (SMR01) and the mental health inpatient and day case datasets (SMR04). Finally, we considered a combined outpatient and admission outcome that included the reporting of three or more separate outpatient appointments (SMR00) plus the reporting of one or more hospital admission (SMR01/SMR04). All outcomes were reported over a 5-year period between 2004-2008.

### Exposure

The main exposure was childhood education and academic ability, relating to the process of learning and educational achievement, especially in educational settings, and the knowledge an individual gains from these educational institutions. We analysed the exposure as a domain (i.e., a group of variables that represent an overarching theme) rather than the individual variables that form its components for two reasons. First, to provide a combined exposure measure that reflects multiple variables in the data rather than performing multiple statistical testing using all of the components in relation to the study outcomes. Second, to conceptualise the components within the wider domain of child education and academic conditions in which the cohort member grew up in.

Variables within the domain included the mean IQ of the school at age 9 (based on Schonell Essential Intelligence Test - Form B [28]), attending a private school (yes/no), type of school attended (primary/secondary), percentage of year absent from school (less than 12.5%/12.5-25.0%/more than 25.0%). Three other variables related to standard school tests, these included the Moray House intelligence test at age 7, a screening tool to identify those who may require specialist education [28]. The Schonell and Adams essential intelligence at age 9, used to screen for poor readers and associated educational difficulties such as dyslexia [28]. Finally, the Moray House verbal reasoning test which measured levels of over or under academic achievement at age 11. This test is often referred to as the ‘eleven plus’ a measurement tool used to assess academic performance and to inform who would go to senior secondary school and who would go to junior secondary school [28,29].

### Childhood confounders

We adjusted for the following confounders chosen based on apriori knowledge [15-18] and recorded in childhood: maternal age, Rutter behaviour (an index of behaviour difficulty), ‘physical grade’ of the cohort member to assess newborns health at birth (good/average/poor/serious), birthweight (below 2500g/over 2500g), maternal occupation (professional/clerical/distribution/skilled/semi-skilled/unskilled/fishwork/manual/no job), father’s social class (I, II, III, IV, V, and unemployed) and sex (male/female).

### Mediators

We explored the role of self-reported mediators in adulthood, selected based on apriori knowledge [10-14] and informed from a directed acyclic graph (DAG), included in Figure 1.

**Figure 1.**
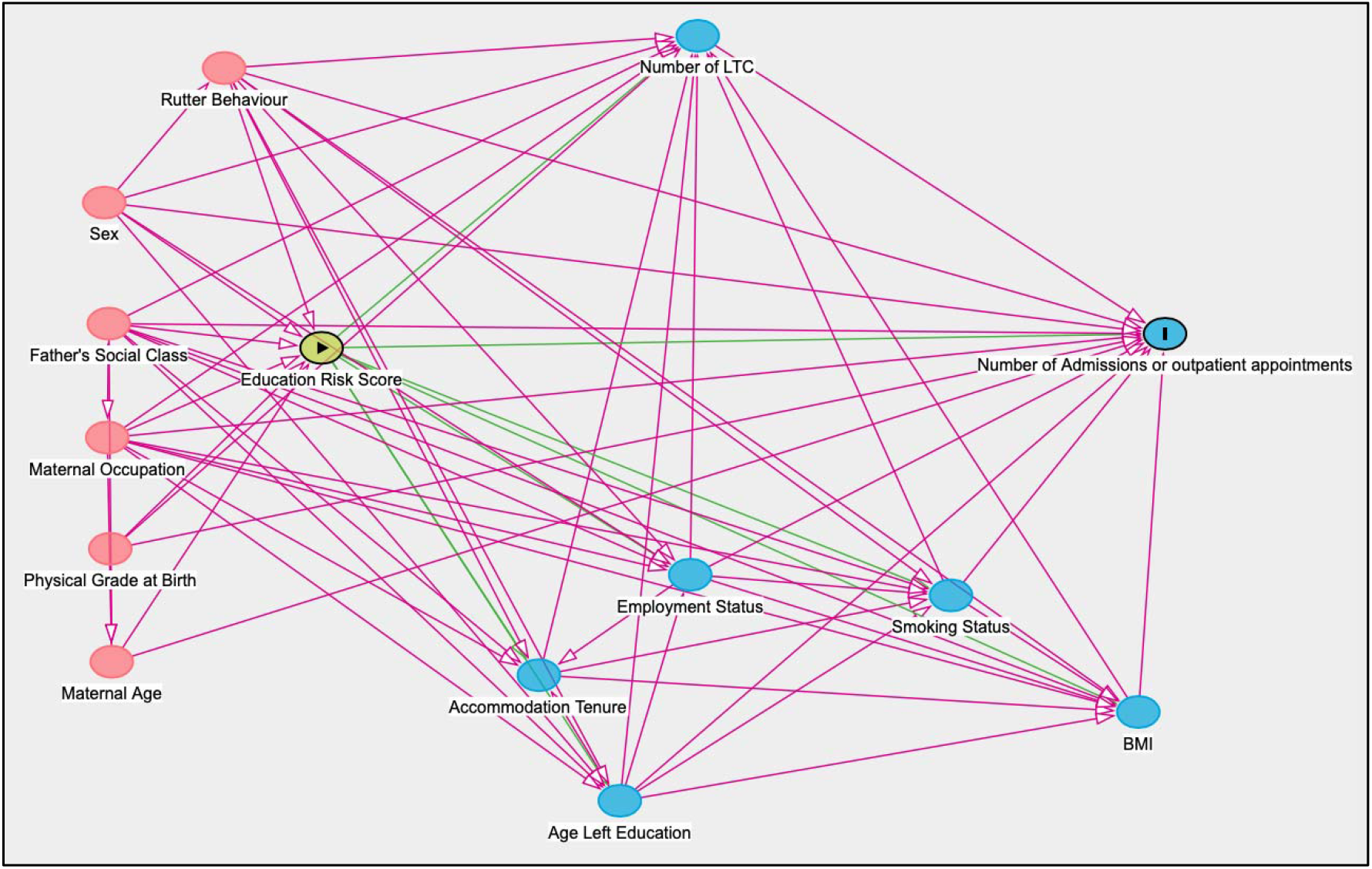
A DAG of the relationship between childhood education and academic ability and hospital appointments and admissions in adulthood.

Mediators measured in the 2001-2003 survey included: body mass index calculated using the following formula - BMI = weight (kg) / height (m)^2^ and included as a continuous measure, smoking status (never/ex-smoker/current smoker), employment status (employed/unemployed/permanently sick or disabled/looking after the family/other), and housing tenure (mortgage/private rent/rent from local authority/other). We considered the mediating role of the presence/number of self-reported LTCs (none, 1 or 2+) reported at the time of the 2001-2003 survey. Self-reported LTCs included: cancer, diabetes, endocrine/metabolic conditions, mental illness, ‘mental handicap’, epilepsy/fits, migraine/headaches, nervous system conditions, cataracts/poor sight, other eye conditions, poor hearing/deafness, ear complaints, stroke, heart attack/angina, hypertension, other heart conditions, varicose veins, other blood vessels/embolic conditions, bronchitis/emphysema, asthma, hayfever, other respiratory conditions, digestive system (ulcer/hernia), upper intestine complaint, complaint of bowel/colon, kidney complaints, bladder conditions, arthritis/rheumatism/fibrositis, back conditions, conditions of bone/joint/muscle, infection/parasitic diseases, skin complaints, and ‘other’ conditions. The final mediator considered was the age the cohort member left school (15 and under/16 and over).

### Analytical sample

The analytical samples included all cohort members who responded to the postal questionnaire conducted between 2001-2003 (n=7183), and our results were based on complete case analysis.

## Statistical analysis

### Step 1: Stepwise backwards elimination to reduce dimensionality of the exposure domain

We performed stepwise backward elimination with admissions and appointments as the outcome, to select variables for inclusion within the education and academic abilities domain. We started with all seven variables, and then variables were removed sequentially if the p-value for a variable exceeded the specified significance level which was set at 0.157. This level was chosen conservatively to reduce the risk of overfitting and is the equivalent to the Akaike information criterion (AIC) [30,31].

### Step 2: Predicted risk scores for each cohort member for each outcome

Logistic regression models were then used to explore the relationship between retained variables and the odds of each of the three outcomes (appointments, admissions and combined appointments and admissions). Based on this logistic regression modelling and using the ‘predict’ function in STATA [32], predicted domain risk scores for each cohort member for each of the three outcomes described in step 1 were calculated, with higher domain risk scores indicating lower education and academic ability. In other words, each cohort member had three predicted score values for the education domain, one for each of the three outcomes, and these predicted risk scores were used correspondingly in step 3. The predicted risk scores for each individual were centred on the mean risk score, and risk scores were bound between -1 and 1.

### Step 3: Multivariable regression analysis including childhood confounders and adult mediators

A series of nested logistic regression models were then used to account for confounders and mediators in the relationship between the education and academic ability domain predicted risk scores and the three outcomes (appointments, admissions, and combined appointments and admissions).

The first model considered the unadjusted relationship between the education and academic ability domain predicted risk scores and the three outcomes, and the second model accounted for childhood confounders. The subsequent six models (models 3-8) included the adult mediators added sequentially into the multivariate model adjusting for childhood confounders. This allowed us to understand the role of mediators for odds of appointments, admissions and combined appointments and admissions, holding all other factors constant.

Regression models were informed by a DAG using DAGitty v3.0 software to explore the most parsimonious possible models (Figure 1). The DAG confirmed the need to include all above variables in the fully adjusted model. All analysis was carried out using STATA version 17 [33], and given that individual academic ability is not entirely independent from school, robust standard errors were calculated and reported.

## Results

### Descriptive results (steps 1-2)

Among 7021 cohort members, 23.9% had ≥5 outpatient appointments, 15.9% had ≥2 hospital admissions and 22.3% had ≥ 3 outpatient appointments plus ≥1 hospital admissions over the five-year period between 2004-2008. The mean number of outpatients appointments over the 5 period was 3.3 (standard deviation 6.8; range 0-117) and the mean number of hospital admission was 0.8 (standard deviation 2.4; range 0-67). The mean age when the cohort members reported 5 outpatient appointments was 53.7 years (standard deviation 1.9; range 49.2-58.8). The mean age was 53.8 years when the cohort members reported 2 hospital admissions (standard deviation 2.1; range 49.2-58.4). The mean age when cohort members reported 3 outpatient appointments plus 1 hospital admissions was 53.8 years (standard deviation 2.0; range 49.2-58.5).

### Regression results (steps 3-4)

In supplementary Tables 1-3 the regression coefficients of outpatient appointments and hospital admissions for the retained variables following stepwise backwards elimination (step 2), and for each of the three outcomes separately are described. Based on these logistic regression models, predicted domain risk scores for each cohort member for each of the three outcomes were calculated.

**Table 1.**
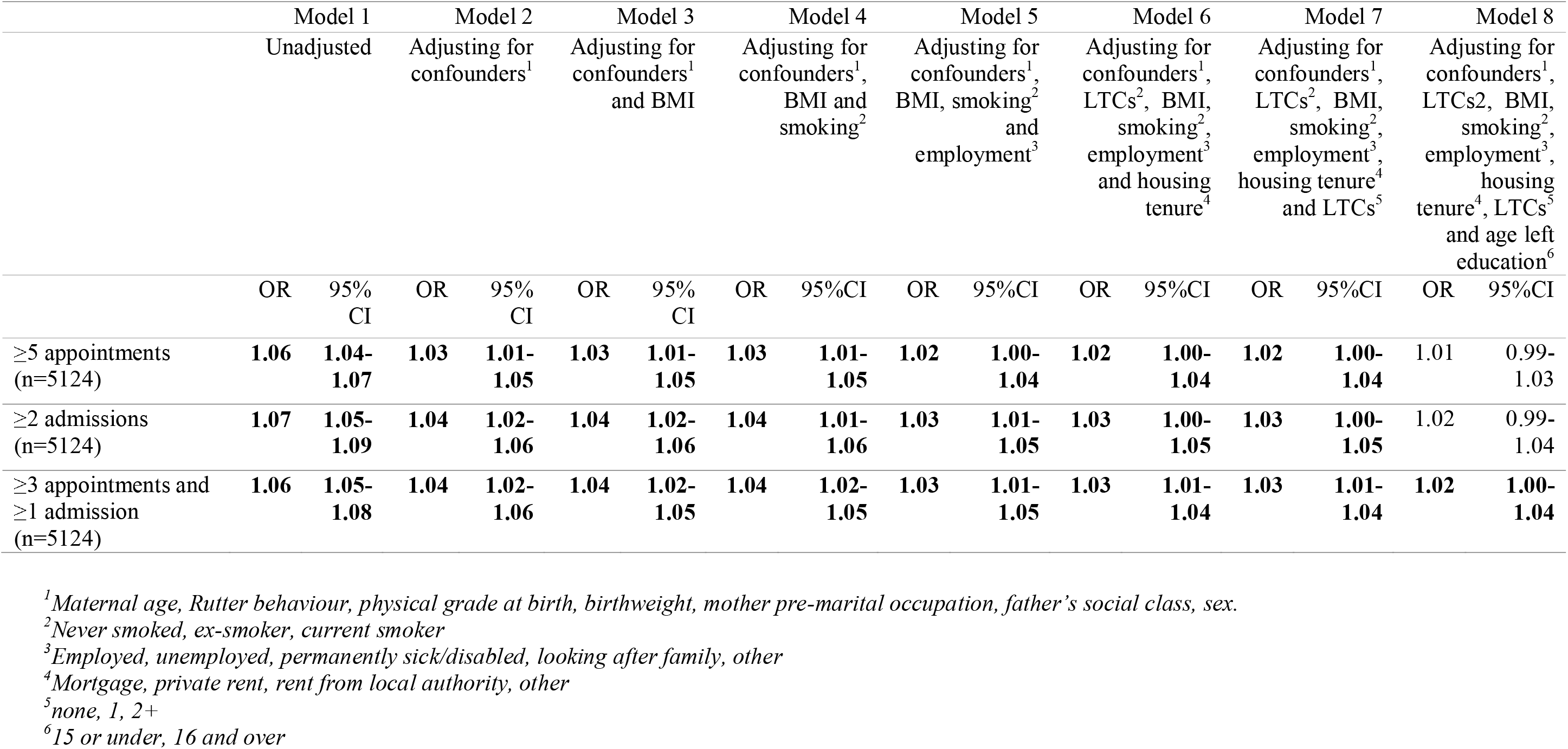
Relationship between education and academic abilities predicted risk score in childhood and secondary care utilisation over a five-year period (2004-2008).

|  | Model 1 |  | Model 2 |  | Model 3 |  | Model 4 |  | Model 5 |  | Model 6 |  | Model 7 |  | Model 8 |  |
| --- | --- | --- | --- | --- | --- | --- | --- | --- | --- | --- | --- | --- | --- | --- | --- | --- |
|  | Unadjusted |  | Adjusting for confounders<sup>1</sup> |  | Adjusting for confounders<sup>1</sup> and BMI |  | Adjusting for confounders<sup>1</sup>, BMI and smoking<sup>2</sup> |  | Adjusting for confounders<sup>1</sup>, BMI, smoking<sup>2</sup> and employment<sup>3</sup> |  | Adjusting for confounders<sup>1</sup>, LTCs<sup>2</sup>, BMI, smoking<sup>2</sup>, employment<sup>3</sup> and housing tenure<sup>4</sup> |  | Adjusting for confounders<sup>1</sup>, LTCs<sup>2</sup>, BMI, smoking<sup>2</sup>, employment<sup>3</sup>, housing tenure<sup>4</sup> and LTCs<sup>5</sup> |  | Adjusting for confounders<sup>1</sup>, LTCs<sup>2</sup>, BMI, smoking<sup>2</sup>, employment<sup>3</sup>, housing tenure<sup>4</sup>, LTCs<sup>5</sup> and age left education<sup>6</sup> |  |
|  | OR | 95% CI | OR | 95% CI | OR | 95% CI | OR | 95% CI | OR | 95% CI | OR | 95% CI | OR | 95% CI | OR | 95% CI |
| ≥5 appointments (n=5124) | **1.06** | **1.04-1.07** | **1.03** | **1.01-1.05** | **1.03** | **1.01-1.05** | **1.03** | **1.01-1.05** | **1.02** | **1.00-1.04** | **1.02** | **1.00-1.04** | **1.02** | **1.00-1.04** | 1.01 | 0.99-1.03 |
| ≥2 admissions (n=5124) | **1.07** | **1.05-1.09** | **1.04** | **1.02-1.06** | **1.04** | **1.02-1.06** | **1.04** | **1.01-1.06** | **1.03** | **1.01-1.05** | **1.03** | **1.00-1.05** | **1.03** | **1.00-1.05** | 1.02 | 0.99-1.04 |
| ≥3 appointments and ≥1 admission (n=5124) | **1.06** | **1.05-1.08** | **1.04** | **1.02-1.06** | **1.04** | **1.02-1.05** | **1.04** | **1.02-1.05** | **1.03** | **1.01-1.05** | **1.03** | **1.01-1.04** | **1.03** | **1.01-1.04** | **1.02** | **1.00-1.04** |

<sup>1</sup>Maternal age, Rutter behaviour, physical grade at birth, birthweight, mother pre-marital occupation, father's social class, sex.
<sup>2</sup>Never smoked, ex-smoker, current smoker
<sup>3</sup>Employed, unemployed, permanently sick/disabled, looking after family, other
<sup>4</sup>Mortgage, private rent, rent from local authority, other
<sup>5</sup>none, 1, 2+
<sup>6</sup>15 or under, 16 and over

Table 1 presents odds ratios of outpatient appointments and hospital admissions as recorded in the health records during the period 2004-2008 in relation to education and academic ability predicted risk scores (step 4) for models adjusting for confounders and then sequentially including mediators - starting with BMI, then smoking status, employment status, housing tenure, presence/number of LTCs and the age the cohort member left education.

In the unadjusted model (model 1), for every one unit increase in the predicted risk score there was a 6% increase in the odds of ≥5 outpatient appointments (OR1.06, 95%CI 1.04-1.07), a 6% increase in the odds of the combined outcome of ≥3 hospital outpatient appointments plus ≥1 admissions (OR1.06, 95%CI 1.05-1.08), and a 7% increase in the odds of ≥2 hospital admissions (OR1.07, 95%CI 1.05-1.09).

Adjusting for childhood confounders (model 2), associations between the exposure and outcomes remained significant but the odds ratios were reduced. For every one unit increase in the predicted risk score there was a 3% increase in the odds of ≥5 outpatient appointments (OR1.03, 95%CI 1.01-1.05), a 4% increase in the odds of ≥3 outpatient appointments plus ≥1 hospital admissions (OR1.04, 95%CI 1.02-1.06), and a 4% increase in the odds of ≥2 hospital admissions (OR1.04, 95%CI 1.02-1.6).

Models 3-8 included adult mediators sequentially into the nested logistic regression models to understand the pathways childhood education and academic ability may affect adult healthcare utilisation. After the inclusion of mediators including BMI, smoking, employment, housing tenure and number of LTCs (models 3-7) the significant relationships identified in the unadjusted model (model 1), and model adjusting for confounders (model 2) were maintained, although the odds ratios were marginally reduced for all three outcomes with the inclusion of each mediator.

After adjusting for the age at which the cohort member left school (model 8), the previously significant relationships between educational and academic ability risk score and ≥5 outpatient appointments (OR1.01, 95%CI 0.99-1.03) and ≥3 hospital admissions (OR1.02, 95%CI 0.99-1.04) were attenuated. However, the relationship between education and academic ability predicted risk scores and ≥3 outpatient appointments plus ≥1 hospital admissions was somewhat attenuated with a reduced effect size but remained statistically significant (OR1.02, 95%CI 1.00-1.04).

## Discussion

In this analysis of a birth cohort born in Scotland, we demonstrated that a domain incorporating factors related to education and academic ability in childhood is related to the number of hospital admissions and outpatient appointments during adulthood. This relationship was not fully explained by the presence of multiple long-term conditions and other potential mediating factors in adulthood. However, the age at which the participant left school substantially mediated this relationship underscoring the significance of lifecourse education on healthcare utilisation.

Our results indicate that the role of education and academic ability in early life on healthcare utilisation maybe independent from factors shaping health in adulthood given the associations were largely explained by including the age at which the participant left school in the models, which is strongly related to academic ability earlier in the lifecourse. To look for potential interventions to address healthcare pressure and patient workload, policies should look beyond known adult risk factors such as unemployment and socioeconomic status [10,14], to wider lifecourse experiences earlier in childhood. In contradiction to what we hypothesised, the associations observed were not fully mediated by adult mediators including the presence/number of LTCs. Instead, the results presented here suggest that the relationship may operate independently as the only mediating factor that had a substantial influence on the effect was the age at which the cohort member left school. Providing further evidence in support of research that has found education to consistently be a key driver in reducing socioeconomic disadvantage, improving health and wellbeing, and promoting health equity [35-37].

We hypothesised that poorer education and academic ability would increase the number of hospital admissions and outpatient appointments. However, the relationship between education and academic ability and health care utilisation is likely to be complex, for example factors related to education (attendance, exclusion and lower educational attainment) have been found to be inversely related to adherence to healthcare attendance and utilisation [38]. Therefore, there is likely to be a pathway through which higher education increases healthcare utilisation through better adherence to healthcare appointments and a better understanding and management of own health, leading however to better health outcomes.

We used three different outcomes to explore different perspectives of healthcare utilisation burden. We found that associations with education and academic ability in childhood were broadly similar across the three outcomes, with marginally more robust associations across the nested models for the combined hospital admissions and outpatient appointments outcome. The combined hospital admissions and outpatient appointments could be hypothesised to represent a more comprehensive conceptualisation of healthcare utilisation given it requires an individual to navigate the time and cost burden of multiple healthcare services and systems.

Research that utilises a lifecourse perspective has consistently demonstrated that the early-life environment can have a significant impact on multiple dimensions of health across the life course [38,39]. However, this research has tended to focus on physical and mental health outcomes in adulthood such as mortality, heart disease, depression, self-rated health, the number of chronic and acute conditions, pain and functional health status [38-42]. Our results suggest that the long arm of childhood might also be relevant for other dimensions of health such as healthcare utilisation independent of the number of chronic health conditions in adulthood.

Our research also supports a body of research that has looked to move away from simple outcomes such as long-term condition counts towards a more sophisticated understanding of health, including an understanding of burdensomeness, work and complexity in the context of MLTCs. A recent qualitative evidence synthesis conducted as part of the MELD-B project described the experiences of people living with multimorbidity[7]. The research identified eight themes of work burden for those living with multimorbidity [7], these themes included learning and adapting; accumulation and complexity; symptoms; emotions; investigation and monitoring; health service and administration; medication; and finance [7]. Our research was able to explore one theme of work burden, however data availability precluded that analysis of all eight themes, and it is important these other themes are incorporated into further analysis.

### Strength and limitations

Utilising ACONF provided in-depth data that allowed us to draw together wider educational determinants of health from early life and measured indicators of burden from link healthcare records in adulthood. This depth of information would not have been available from most electronic health care records in either primary or secondary care. Further, utilising data recorded at various timepoints allowed for the temporal ordering of the variables to be established.

Despite this, the small sample size of participants with any one LTC precluded the opportunity to investigate subgroup analysis, exploring how different clusters of conditions may have mediated the relationships considered differently. This is particularly important given that some LTCs are likely to be more challenging than others for patients in terms of symptoms, impacting self-management demands (burden of treatment) and health-related quality of life [43-47]. The small sample size also restricted the dimension of burden we could consider. Holland et al., [7] identified eight themes of work burden for those living with multimorbidity, however our research could only partially capture one of these themes of work – ‘health service and administration’. Further research should continue to explore alternative datasets that capture both the early life course and a deeper understand of burden, particular for those living with multimorbidity.

The traced cohort members (in 1999) were more likely to be women, from a higher birth social class and have a higher childhood cognition score than non-responders [28], all factors that might also be related to reduced healthcare utilisation in adulthood. This means we might be underestimating the prevalence of hospital admissions and outpatient appointments. We could not adjust for important health behaviours such as diet and physical activity as the data were not available. Also, we did not account for the specific causes for the outpatient appointments or hospital admissions due to data limitations including data quality and high levels of missing data particularly ICD10 condition codes within the outpatient dataset (SMR00).

We acknowledged in the methods section that our outcome cut-off for healthcare utilisation burden was arbitrary, and other researchers may propose alternative cut-offs. However, the research team chose a cut-off we felt may represent burden at a population level, and this cut off was agreed with the support of members of our patient and public advisory board. However, we were unable to considered other aspects of health services utilisation burden our patient and public advisory board felt may be important such as general practice appointments and social services appointments.

We were limited to the variables collected at the time of the survey and as such some of the variables are likely to now be outdated. In particular, the three standardised school tests used widely at the time of the survey in Scotland are not in use today. The school tests were also designed to identify those who performed poorly in academic tests or required additional education support. As such, the Scottish education system of the time encouraged the screening and segregation of some pupils, and this may have resulted in the widening of educational disparities and inequalities.

It is also important to note that we were unable to account for health literacy, which is distinct from literacy and is a personal attribute defined as the ability to access, understand, and use health-related information [47,48]. This may have been an important omission given the significant literature around the relationship between health literacy and healthcare access and utilisation [50-52].

Finally, the ACONF cohorts have only been interviewed once during adulthood. As a result, the mediators and outcomes considered in this paper relate to a period around 20 years ago. Future sweeps of ACONF data collection are being planned, and when available we recommend repeating the analysis presented here with the latest sweeps of data to explore if the associations described are maintained across the adult lifecourse.

## Conclusion

Education and academic ability in early life may be related to the burden of multiple hospital admissions and outpatient appointments later in life with this relationship not be being fully explained by the presence of multiple long-term conditions and other factors in adulthood. However, the age at which the participant left school seems to mediate this relationship underscoring the significance of lifecourse education on health. Specific interventions may look to support children to stay in school and optimise their ability to learn and develop, recognising that some will need additional support that takes account of their specific circumstances. Our results also go some way to suggest that the burden of healthcare utilisation extends beyond known factors in adulthood and can be linked back to factors earlier in the lifecourse. This is important to inform early life interventions toward better long-term health and reduce healthcare burden.

## Conflict of Interest

R.O. is a member of the National Institute for Health and Care Excellence (NICE) Technology Appraisal Committee, member of the NICE Decision Support Unit (DSU), and associate member of the NICE Technical Support Unit (TSU). She has served as a paid consultant to the pharmaceutical industry and international reimbursement agencies, providing unrelated methodological advice. She reports teaching fees from the Association of British Pharmaceutical Industry (ABPI).

## Author Contributions

S.F., N.A., S.P., R.O., S.S. and A.B. contributed to the conceptualisation of the MELD-B project. S.S., N.A., and S.F. obtained the datasets. All authors contributed to the conceptualisation of the paper. S.S., and N.A. led the design and planning of the paper. S.F., N.A., S.P., R.O., S.S. and A.B. supported the design, planning and reviewing of the statistical analysis plan. S.S. performed the statistical analysis. S.S. prepared all figures and graphs. S.S., and N.A. produced the initial draft of the manuscript. All authors were involved in editing and reviewing the manuscript, and approved the final manuscript. S.S., N.A., and S.F. take responsibility for the data and research governance.

## Data Availability Statement

Anonymised data can be requested by accredited researchers with appropriate research governance and ethics approvals via the Grampian Data Safe Haven (https://www.abdn.ac.uk/research/digital-research/dash.php).

## Acknowledgement

We would like to acknowledge all other members of the MELD-B Consortium, and we thank the participants of the ACONF study.

## Funding Statement

This study is independent research funded by the National Institute for Health and Care Research (NIHR) Artificial Intelligence for Multiple Long-Term Conditions (AIM) ‘Multidisciplinary Ecosystem to study Lifecourse Determinants and Prevention of Early-onset Burdensome Multimorbidity (MELD-B)’, (reference number NIHR203988). The views expressed are those of the authors and not necessarily those of the NHS, NIHR or the Department of Health and Social Care.

## Notes

### Author Declarations

Ethics approval for the MELD-B project has been obtained from the University of Southampton Faculty of Medicine Ethics committee (ERGO II Reference 66810).

