## Supplementary Tables 1-3 for "Exploring the Relationship Between Education and Academic Ability in Childhood with Healthcare Utilisation in Adulthood: Findings from the Aberdeen Children of the 1950’s (ACONF)"

*Table 1. Coefficients from logistic regression modelling of 5 or more outpatient appointments. Variables retained following stepwise backwards elimination. (n=5124).*

|  |  | Coef | 95% CI |
| --- | --- | --- | --- |
| School type | Primary | Ref | Ref |
|  | Secondary | 1.14 | 0.99-1.31 |
| School mean IQ |  | 0.98 | 0.97-0.99 |
| Schonell and Adams Essential intelligence test score |  | 0.99 | 0.98-0.99 |

*Table 2. Coefficients from logistic regression modelling of 2 or more hospital admissions. Variables retained following stepwise backwards elimination. (n=5124)*

|  |  | Coef | 95% CI |
| --- | --- | --- | --- |
| Private school | No | Ref | Ref |
|  | Yes | 0.33 | 0.07-1.37 |
| Absence from school | 12.5% or less | Ref | Ref |
|  | 12.5-25.0% | 1.54 | 0.93-2.54 |
|  | 25.0% or over | 2.56 | 0.46-14.3 |
| Moray house intelligence test score |  | 0.99 | 0.98-0.99 |
| Schonell and Adams Essential intelligence test score |  | 0.99 | 0.98-0.99 |

|  |  | Coef | 95% CI |
| --- | --- | --- | --- |
| School IQ |  | 0.98 | 0.97-0.99 |
| Schonell and Adams Essential intelligence test score |  | 0.98 | 0.98-0.99 |

*Table 3. Coefficients from logistic regression modelling of 3 or more outpatient appointment and 1 or more hospital admissions. Variables retained following stepwise backwards elimination. (n=5124)*
